# Predicting 30-Day Heart Failure Readmissions Using Machine Learning: Insights From the Kansas Health Information Network (KHIN)

**DOI:** 10.64898/2026.05.18.26353537

**Authors:** Minjung Kim, Jun Yan, Jason Wasfy, Robert H. Aseltine

## Abstract

**Background:** Heart failure (HF) is a major contributor to inpatient hospital utilization, with persistently high 30-day readmission rates. Existing prediction tools are frequently restricted to primary-diagnosis HF admissions, potentially excluding clinically relevant HF-related hospitalizations.

**Objectives:** To develop and validate risk prediction models using machine learning (ML)-based risk prediction models to predict 30-day readmissions among patients with HF using the Kansas Health Information Network, a statewide health information exchange.

**Methods:** This retrospective cohort study analyzed HF hospitalizations using predictors including demographics, comorbidities, laboratory results, medications, clinical quality metrics for diabetes and kidney disease management, and prior healthcare utilization. Five ML models, including regularized logistic regression, random forest, extreme gradient boosting, categorical boosting, and deep neural network, were trained using stratified 5-fold cross-validation. Model performance was evaluated on an independent test set using the area under the receiver operating characteristic curve (AUROC), area under the precision-recall curve (AUPRC), misclassification rate (MCR), and Brier score.

**Results:** Among 2,734 HF patients, the 30-day readmission rate was 27%. The XGBoost model achieved the best discrimination (AUROC=0.75; AUPRC=0.58; MCR=0.21). Patients in the highest-risk decile had a positive predictive value of 76%, accounted for approximately one-third of all 30-day readmissions, and had a 3.3-fold enrichment compared with baseline risk. The key predictors included prior hospital utilization, diabetes and kidney disease management indicators, and comorbidity burden.

**Conclusions:** Risk stratification using routinely collected clinical data identified a subgroup at elevated risk for 30-day readmission. These findings support the potential role of data-driven risk prediction to inform targeted transitional care.

**CLINICAL PERSPECTIVES:** *What is New?:* This study is among the first to utilize health information exchange data to develop machine learning models for predicting 30-day heart failure readmissions, including patients with HF as either a primary or secondary diagnosis. Unlike prior models restricted to primary HF admissions, this approach includes hospitalizations in any diagnosis position, reflecting real-world acute-care utilization. Notably, our machine learning model correctly identified approximately three in four patients classified as high-risk in the top decile of predicted scores, highlighting its ability to identify individuals at greatest likelihood of early readmission

*What are the Clinical Implications?:* Accurately identifying a group of high-risk patients has meaningful implications for care delivery. By correctly capturing actual readmissions within a small, prioritized segment of the population, ML-driven risk stratification can guide clinicians and care management teams in targeting transitional care resources where they are most needed. Such focused intervention strategies may allow for efficient targeting of limited health care resources, improve post-discharge outcomes, enhance operational efficiency, and potentially reduce preventable HF readmissions.

## INTRODUCTION

Heart failure (HF) remains a significant public health challenge, affecting approximately 6.7 million adults in the United States as of 2023, with projections estimating an increase to 8 million by 2030. It is a leading cause of hospitalization, accounting for over 1 million hospital admissions annually and contributing to over $30 billion in healthcare costs each year.^1^ Among hospitalized HF patients, 30-day readmission rates range from 17% to 24%, making HF a key focus for hospital-based interventions^2,3^. In response, the Centers for Medicare & Medicaid Services (CMS) incorporated HF into the Hospital Readmissions Reduction Program (HRRP) in 2012, implementing financial penalties for hospitals with excessive readmissions^4,5^. While the HRRP has contributed to modest reductions in readmission rates, risk-adjusted 30-day readmissions after HF hospitalization declined by roughly 1% per year around the time of HRRP implementation and then stabilized^3,6,7^. Much of the apparent decrease in risk-standardized readmissions after HRRP appear related to increased coding of comorbidities, which increase the expected readmission rate^8^.

Current strategies to reduce HF readmissions primarily target patients with HF as the principal diagnosis, often overlooking those with HF as a secondary diagnosis. Although the HRRP targets only hospitalizations with a primary HF diagnosis, admissions with HF coded as a secondary diagnosis remain excluded from most risk stratification frameworks, despite evidence suggesting a comparable risk of readmission. The patient hospitalized with HF as the secondary diagnosis, which frequently presents with complex comorbidities, remains underrepresented in research despite evidence suggesting a comparable risk of readmission^7,9,10^. The lack of comprehensive predictive models encompassing this broader patient population limits clinicians’ ability to identify and mitigate readmission risks effectively.

Recent studies predicting 30-day readmission risk for HF patients, whether admitted with HF as a primary or secondary diagnosis, highlight the evolving landscape of predictive modeling. Traditional models such as the LACE index and HOSPITAL score demonstrate moderate predictive ability but lack adaptability to individual patient characteristics^11–14^. These limitations have prompted the introduction of machine learning (ML) models which have shown variable improvements over logistic regression approaches in structured electronic health records (EHR) data, achieving AUC values between 0.72 and 0.79^15–18^. Biomarker-based models, incorporating NT-pro BNP and point-of-care ultrasound (POCUS) assessments, have further enhanced predictive accuracy^19–24^. Despite their potential, these approaches rely on specialized biomarkers or imaging diagnostics that require additional tests and workflow requirements, which may limit widespread implementation across healthcare settings. Recent studies underscore the importance of integrating a broad range of predictive factors to improve HF readmission risk stratification. Key predictors of readmission include clinical factors (e.g., comorbidities such as chronic kidney disease and diabetes, prior hospitalizations, and biomarker levels), as well as demographic and social determinants (e.g., age, socioeconomic status, and healthcare access)^25–29^.

This study addresses these gaps by using ML models to predict 30-day readmission among hospitalizations with HF present among diagnosis codes, without restricting analyses to primary-diagnosis definitions. Using a rich dataset from the Kansas Health Information Network (KHIN), the research incorporates diverse predictors, including patient demographics, clinical quality measures, prior healthcare utilization, and admission characteristics. By leveraging routinely collected structured data, this study provides actionable insights to optimize post-discharge strategies, prioritize scarce clinical resources to help patients avoid preventable readmission, and potentially reduce HF readmissions.

## METHODS

### Data and Cohort Definition

This retrospective cohort study utilized EHR from the Kansas Health Information Network (KHIN), one of the most mature Health Information Exchange (HIE) networks in the United States. KHIN compiles data from approximately 500-600 healthcare facilities across diverse clinical settings, healthcare providers, and electronic health record platforms.

HIE data remain underutilized in research due to challenges such as mixed data formats (e.g., varying coding schemes) and incomplete entries^30,31^. Despite these limitations, prior studies have demonstrated their utility for large-scale outcomes research and care quality improvement^32,33^.

KHIN data included inpatient and outpatient information from facilities across Kansas and neighboring states. The dataset consisted of patient demographics, encounter details, diagnosis codes, procedure codes, laboratory orders and results, and medication records, offering a rich source of information for this analysis.

This retrospective cohort study focused on patients diagnosed with HF who had hospital admissions during the observation period from January 1, 2016, to January 31, 2018. The index admission was observed between January 1, 2016, and December 31, 2017, to assess whether a 30-day readmission occurred after the initial hospitalization. Hospitalizations were defined as encounters with a length of stay greater than zero days and included both inpatient and observation admissions to capture clinically meaningful acute-care utilization across care settings. Because diagnosis position (primary vs secondary) was not consistently available in KHIN, hospitalizations were identified based on the presence of HF diagnosis codes regardless of hierarchy.

To enhance predictor construction, an additional observation period from January 1, 2015, to December 31, 2015, was used to measure clinical quality and assess prior healthcare utilization, including the number of emergency room (ER) visits, hospital admissions, and 30-day readmissions occurring 1 year preceding the index admission. Predictors were derived solely from data available up to discharge from the index admission.

Eligible patients had records with a positive length of stay, at least one diagnosis, one medication entry, and one laboratory result during the observation period. The index admission was defined as the first hospital visit during the 2016–2017 observation period for each unique HF patient. Supplemental Figure 1 displays a flowchart of the study design, outlining patient inclusion and exclusion criteria and illustrating the cohort selection process. The study then tracked 30-day readmission events at any hospital in the KHIN network following the index admission to identify patterns and predictors of readmission risk.

### Demographics

Self-reported demographic information included age and sex (female and male). The age at the index admission was recorded for each patient.

### Diagnosis Data

Diagnosis information was collected for all medical encounters during the 1 year prior to the index admission, including the index admission itself. This information, encoded using ICD-9 and ICD-10 codes in KHIN, was normalized for consistency. ICD-9 codes were converted to ICD-10 codes using the R package “touch.” The diagnostic codes were aggregated by their first three digits.

### Medication Data

Medication data were recorded using RX codes (93%) and NDC codes (7%). To normalize these two types of medication data, all codes were converted to Anatomical Therapeutic Chemical (ATC) codes, facilitating interpretation by therapeutic categories^34^.

### Clinical Metrics for Diabetes and Kidney Function

Clinical metrics related to diabetes and kidney function were derived from diagnostic and laboratory data. Diabetes-related variables were categorized using the quality measure of Hemoglobin A1c (HbA1c) Poor Control (>9%) and associated diagnosis codes (E08, E09, E10, E11, E13). For kidney function, estimated glomerular filtration rate (GFR) thresholds defined three categories: normal (GFR ≥ 60), kidney disease (GFR < 60), and kidney failure (GFR < 15), along with diagnosis codes (N17, N18, N19). Composite variables reflected the management quality for both conditions, incorporating laboratory results and diagnosis history. The clinical metrics for diabetes and kidney disease management variables are categorized as good, bad, or warning. The details of these new metrics are provided in Supplemental Table 1.

### Hospital Utilization Prior to Index Admission

Hospital utilization one year before index admissions included the number of emergency room (ER) visits, the number of hospital admissions, and the number of 30-day readmissions.

### Machine Learning Models

The prediction of 30-day readmission was framed as a supervised classification problem, and several ML models were utilized (Figure 2). Ensemble learning methods included Random Forest (RF)^35,36^, Extreme Gradient Boosting (XGBoost)^37,38^, and CatBoost^39,40^. In addition, Regularized Logistic Regression (LR)^41^ was included as a benchmark model due to its simplicity and intrinsic interpretability. A Deep Neural Network (DNN)^42^ was also used, consisting of fully connected layers with ReLU activation, optimized using the Adam optimizer, with dropout, and early stopping to reduce overfitting.

**Figure 1.**
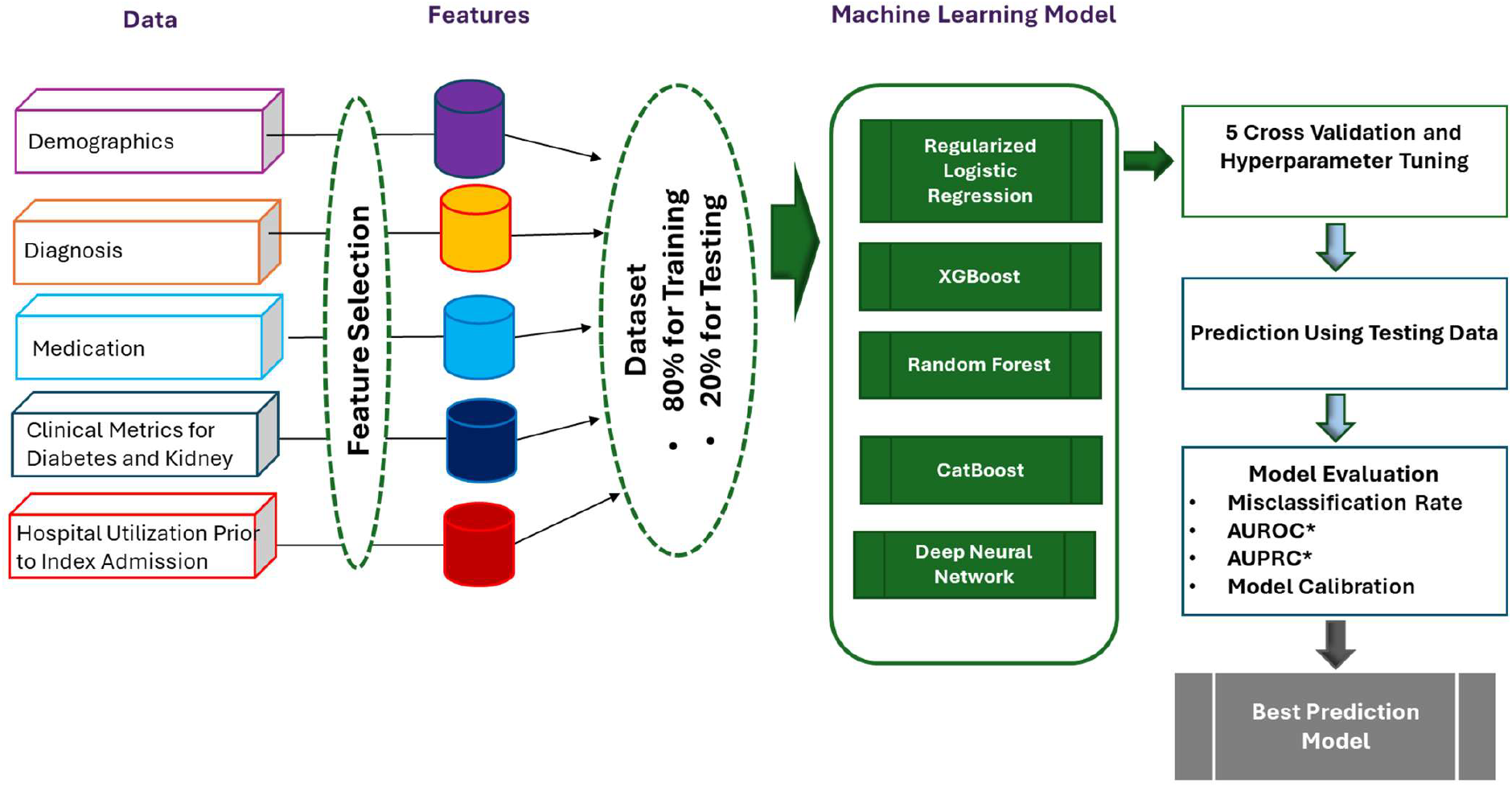
Workflow of Machine Learning–Based Prediction of 30-Day Heart Failure Readmission: Overview of the analytic workflow, including cohort selection from the Kansas Health Information Network, feature construction from demographics, comorbidities, laboratory values, medications, clinical quality measures, and prior healthcare utilization, model development using multiple machine learning algorithms, and evaluation on an independent testing dataset. Model performance was assessed using discrimination, calibration, and clinically actionable threshold-based metrics. **AUROC*:** Area Under the Receiver Operating Characteristic Curve; **AUPRC*:** Area Under the Precision-Recall Curve.

**Figure 2.**
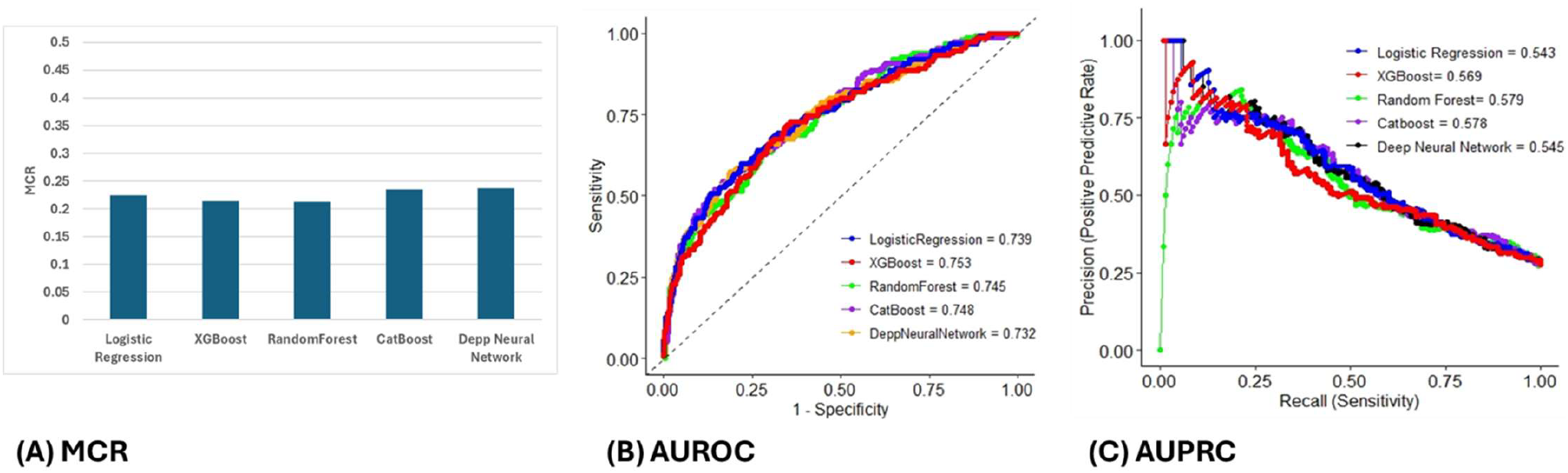
Performance of Machine Learning Models for Predicting 30-Day Readmission: Performance of five machine learning models evaluated on the independent testing dataset. Discrimination was assessed using the area under the receiver operating characteristic curve (AUROC) and the area under the precision–recall curve (AUPRC), and overall classification error was summarized by the misclassification rate (MCR). XGBoost and RF models demonstrated the strongest overall performance across discrimination, classification accuracy, and clinically relevant operating thresholds.

The dataset was divided into training (80%) and test (20%) sets using stratified random sampling ensuring consistent outcome prevalence across both sets. Model training involved five-fold cross-validation to prevent overfitting, and hyperparameter tuning was conducted using a grid-search strategy, with an emphasis on maximizing the area under the receiver operating characteristic curve (AUROC).

To address imbalance, stratified sampling and class-weighting scheme were applied, assigning weights inversely proportional to class frequencies. Model performance was evaluated using multiple metrics, including AUROC, area under the precision-recall curve (AUPRC), and misclassification rate (MCR), to provide a balanced assessment of discrimination and classification performance across models.

### Model Evaluation

Model performance was assessed on an independent test set using both threshold-independent and threshold-dependent metrics. Threshold-independent metrics included the AUROC, AUPRC, MCR, and model calibration using the Brier Score. In addition to these global performance indicators, we evaluated sensitivity and positive predictive value (PPV) at fixed specificity thresholds of 90% and 95%. Together, these metrics provided a balanced assessment of discrimination, calibration, and clinically actionable performance for predicting 30-day readmissions.

## RESULTS

### Predictors of 30-day readmission

A total of 2,734 patients admitted with a diagnosis of heart failure (HF) were included in the study (Table 1). The overall 30-day readmission rate was 27%. Readmitted patients were more likely to be younger (age <65), to have diastolic or unspecified HF. They also demonstrated a higher burden of comorbidities, including poorly controlled diabetes and kidney disease, longer length of stay (LOS), and greater healthcare utilization in the year preceding the index admission (including emergency department visits, hospital admissions, and prior 30-day readmissions).

**Table 1.** Baseline Characteristics of Heart Failure Patients With and Without 30-Day Readmission: Baseline demographic characteristics, heart failure subtypes, clinical quality measures, length of stay, and healthcare utilization in the year prior to the index admission are shown for patients with and without 30-day readmission. Continuous variables are presented as median [interquartile range], and categorical variables are presented as number (percentage). p_values reflect univariate comparisons between groups.

| Variable |  | Without 30-Day Readmission | With 30-Day Readmission | p_value |
| --- | --- | --- | --- | --- |
| Number of Observations: n (%) |  | 1987 (73%) | 747 (27%) |  |
| Demographics | AGE (Median, [IQR*]) | 60, [54-64] | 61, [54-65] | 0.087 |
|  | Female | 946 (48%) | 357 (48%) | 0.967 |
| HF Subtype | Systolic HF | 387 (19%) | 130 (17%) | 0.238 |
|  | Diastolic HF | 294 (15%) | 124 (17%) | 0.268 |
|  | Combined HF | 80 (4%) | 31 (4%) | 0.970 |
|  | Unspecified HF | 1177 (59%) | 467 (63%) | 0.129 |
|  | Other HF | 188 (9%) | 56 (7%) | 0.006 |
| Clinical Quality Measure | Diabetes Good Management | 969 (49%) | 290 (39%) | <0.001 |
|  | Diabetes Bad Management | 857 (43%) | 387 (52%) | <0.001 |
|  | Diabetes Management Warning | 161 (8%) | 70 (9%) | 0.324 |
|  | Kidney Disease Good Management | 942 (47%) | 209 (28%) | <0.001 |
|  | Kidney Disease Bad Management | 237 (12%) | 128 (17%) | <0.001 |
|  | Kidney Disease Management Warning | 808 (41%) | 410 (55%) | <0.001 |
| Hospital Utilization | Admission Via Index | 220 (11%) | 89 (12%) | 0.581 |
|  | Length of Stay (Median, [IQR*]) | 3, [2-5] | 4, [2-7] | 0.014 |
|  | N_30Day* (Median, [IQR*]) | 0, [0-0] | 0, [0-1] | 0.004 |
|  | 0 | 1649 (83%) | 560 (75%) | <0.001 |
|  | 1 | 159 (8%) | 52 (7%) |  |
|  | 2 ≤ N_30Day* ≤ 3 | 119 (6%) | 75 (10%) |  |
|  | N_30Day* ≥ 4 | 60 (3%) | 60 (8%) |  |
|  | N_AD* (Median, [IQR*]) | 0, [0-0] | 0, [0-1] | <0.001 |
|  | 0 | 1391 (70%) | 359 (48%) | <0.001 |
|  | 1 ≤ N_AD* ≤ 2 | 358 (18%) | 156 (21%) |  |
|  | 3 ≤ N_AD* ≤ 5 | 159 (8%) | 127 (17%) |  |
|  | N_AD* ≥ 6 | 79 (4%) | 105 (14%) |  |
|  | N_ER* (Median, [IQR*]) | 0, [0-0] | 0, [0-0] | 0.004 |
|  | 0 | 1649 (83%) | 560 (75%) | <0.001 |
|  | 1 | 159 (8%) | 60 (8%) |  |
|  | 2 ≤ N_ER* ≤ 3 | 119 (6%) | 75 (10%) |  |
|  | N_ER ≥ 4 | 60 (3%) | 52 (7%) |  |

IQR\*: Inter-Quartile Range; N\_30Day: Number of 30Day Readmissions One Year before Index Admission; N\_AD: Number of Admissions One Year before Index Admission, N\_ER: Number of ER Visits One Year before Index Admission.

**Table 2.** Performance of the XGBoost Model Stratified by Sex, Age Group, and HF Subtype: Discriminative performance of the XGBoost model across demographic (sex, age group) and clinical (heart failure subtype) subgroups in the testing dataset. Metrics include area under the receiver operating characteristic curve (AUROC), area under the precision–recall curve (AUPRC), sensitivity, and positive predictive value evaluated at fixed specificity thresholds of 90% and 95%.

| Subgroup | AUROC | AUPRC | Accuracy | Sensitivity |  | Positive Predictive Value |  |
| --- | --- | --- | --- | --- | --- | --- | --- |
|  |  |  |  | 90% Specificity | 95% Specificity | 90% Specificity | 95% Specificity |
| Male | 0.754 | 0.632 | 0.798 | 0.506 | 0.367 | 0.667 | 0.744 |
| Female | 0.750 | 0.525 | 0.756 | 0.437 | 0.310 | 0.620 | 0.688 |
| 18≤Age<45 | 0.810 | 0.746 | 0.846 | 0.600 | 0.500 | 0.667 | 0.833 |
| 45≤Age<65 | 0.737 | 0.533 | 0.773 | 0.429 | 0.297 | 0.591 | 0.659 |
| Age≥65 | 0.762 | 0.644 | 0.770 | 0.469 | 0.388 | 0.697 | 0.792 |
| Systolic HF | 0.760 | 0.643 | 0.823 | 0.500 | 0.455 | 0.611 | 0.714 |
| Combined HF | 0.794 | 0.779 | 0.821 | 0.600 | 0.500 | 0.750 | 0.833 |
| Diastolic HF | 0.751 | 0.572 | 0.792 | 0.524 | 0.381 | 0.688 | 0.727 |
| Unspecified HF | 0.743 | 0.598 | 0.749 | 0.410 | 0.330 | 0.641 | 0.733 |

**AUROC\***: Area Under the Receiver Operating Characteristic Curve; **AUPRC\***: Area Under the Precision-Recall Curve

**Table 3.** Comparison of Feature Prevalence Between True Positive and False Positive Patients in the High-Risk Decile Group Identified by the XGBoost Model: Prevalence of the 25 most common features among true positive (n = 42) and false positive (n = 13) patients within the highest-risk decile of predicted readmission probability identified by the XGBoost model. Values are presented as number (percentage). This analysis is descriptive and intended to characterize clinical and utilization patterns associated with correct and incorrect classification within the high-risk group.

| Features | True Positive (n = 42) | False Positive (n = 13) |
| --- | --- | --- |
| Long term (current) drug therapy | 42 (100%) | 13 (100%) |
| Admission before index admission | 40 (95%) | 12 (92%) |
| Type 2 diabetes mellitus | 36 (86%) | 8 (62%) |
| Other disorders of fluid, electrolyte and acid-base balance | 36 (86%) | 9 (69%) |
| Disorders of lipoprotein metabolism and other lipidemia | 33 (79%) | 10 (77%) |
| Sleep disorders | 32 (76%) | 4 (31%) |
| Overweight and obesity | 32 (76%) | 3 (23%) |
| Personal history of other diseases and conditions | 32 (76%) | 9 (69%) |
| Personal history of certain other diseases | 32 (76%) | 9 (69%) |
| Body mass index | 32 (76%) | 7 (54%) |
| Major depressive disorder, single episode | 31 (74%) | 6 (46%) |
| Kidney Disease Management Warning | 31 (74%) | 11 (85%) |
| Chronic ischemic heart disease | 30 (71%) | 7 (54%) |
| Abnormalities of breathing | 30 (71%) | 4 (31%) |
| Chronic kidney disease | 30 (71%) | 8 (62%) |
| Other anxiety disorders | 30 (71%) | 6 (46%) |
| Gastro-esophageal reflux disease | 30 (71%) | 9 (69%) |
| Acute kidney failure | 29 (69%) | 10 (77%) |
| Other anemias | 28 (67%) | 7 (54%) |
| Other disorders of urinary system | 28 (67%) | 9 (69%) |
| Dependence on enabling machines and devices, necrotizing enterocolitis | 28 (67%) | 10 (77%) |
| Unspecified HF | 28 (67%) | 9 (69%) |
| Malaise and fatigue | 27 (64%) | 5 (38%) |
| Respiratory failure, not elsewhere classified | 27 (64%) | 7 (54%) |
| Diabetes Management Warning | 27 (64%) | 8 (62%) |

### Predictive Performance of ML Models

The model evaluation on the testing dataset closely aligned with the cross-validation results obtained during training, supporting the models’ generalization capability (Figure 2). Training performance and variability across five-fold cross-validation are additionally summarized in Supplemental Figure 2. Among the ensemble methods, XGBoost and RF demonstrated consistent performance, each achieving a MCR of 21%, AUROC of 0.75, and AUPRC ranging from 0.57 to 0.58. At a fixed specificity of 90%, these models achieved sensitivity values between 0.44 and 0.46, coupled with a positive predictive value (PPV) of 0.62 to 0.63. When the specificity threshold was raised to 95%, sensitivity decreased to a range of 0.34 to 0.35, while PPV increased to 0.71 to 0.72. CatBoost followed with an MCR of 24%, an AUROC of 0.75, and an AUPRC of 0.58.

Model calibration, an essential aspect of predictive model evaluation, was further assessed using the Brier Score, which quantifies the accuracy of probabilistic predictions. The best performance was observed by XGBoost (0.1600) and RF (0.1645), followed by regularized LR (0.2024). The DNN and CatBoost models showed weaker calibration (0.2244 and 0.2488, respectively). Compared with the null-model Brier score (0.197) for a cohort with 27% event prevalence, CatBoost, regularized logistic regression, and DNN exhibited higher Brier scores, indicating worse probabilistic accuracy than the naive baseline. Although multiple models demonstrated comparable discriminative performance, XGBoost achieved the best overall balance of discrimination and calibration and was therefore selected as the primary predictive model for subsequent analyses. Additional calibration assessment using decile reliability plots and evaluation of clinical utility using decision curve analysis are presented in Supplemental Figure 3. These analyses demonstrated acceptable calibration and favorable net clinical benefit across clinically relevant risk thresholds.

### Subgroup Performance Analysis

Model performance was further examined across key clinical and demographic subgroups, including sex (male vs. female), age category (18–44, 45–64, ≥65 years), and HF subtype (SHF, CHF, DHF, UHF) to evaluate the stability of model discrimination and predictive utility across clinically relevant patient strata. Subgroup analyses showed that model performance remained stable across sex, age strata, and HF subtypes.

Discrimination was similar for females (AUROC = 0.754; AUPRC = 0.632) and males (AUROC = 0.750; AUPRC = 0.525). Model performance varied modestly by age group, with the highest discrimination observed among patients aged 18–44 (AUROC = 0.810; AUPRC = 0.747). Across HF subtypes, discrimination was highest for combined HF (CHF; AUROC = 0.794; AUPRC = 0.779). The positive predictive value remained consistently high at 95% specificity (0.659–0.833) across all groups, underscoring the model’s reliability in identifying patients at truly elevated readmission risk under conditions of high specificity. Overall, no subgroup exhibited evidence of performance decline, indicating equitable model performance across demographic and clinical categories.

### Permutation Feature Importance

Permutation Feature Importance was used to quantify the contribution of each predictor to the model’s predictive performance. For each feature, values were randomly permuted across observations while holding all other variables constant, and the resulting decline in model performance (1-AUROC) was calculated across 10 permutations for each feature. We included 467 features identified as significant predictors of 30-day readmission, with the mean importance values computed for each variable across all ML models.

Figure 3 highlights the top 25 features with the highest importance scores in XGBoost model. Comparing the top 25 features of RF with other ML models (Supplemental Figure 4), the results reveal consistency in the most important predictors across models, with only minor variations in rank order. The top five predictors consistently included the number of admissions, and the number of 30-day readmissions in the year preceding the index admission, age, diagnosis of “chronic kidney disease”, “personal risk factors, not elsewhere classified”, kidney disease management. Additional highly ranked predictors included the diagnosis of “disorder of mineral metabolism”, “malaise and fatigue”, and “overweight and obesity.” A comprehensive comparison of relative permutation feature importance of the top 25 features across the five ML models is presented in Supplemental Table 3.

**Figure 3.**
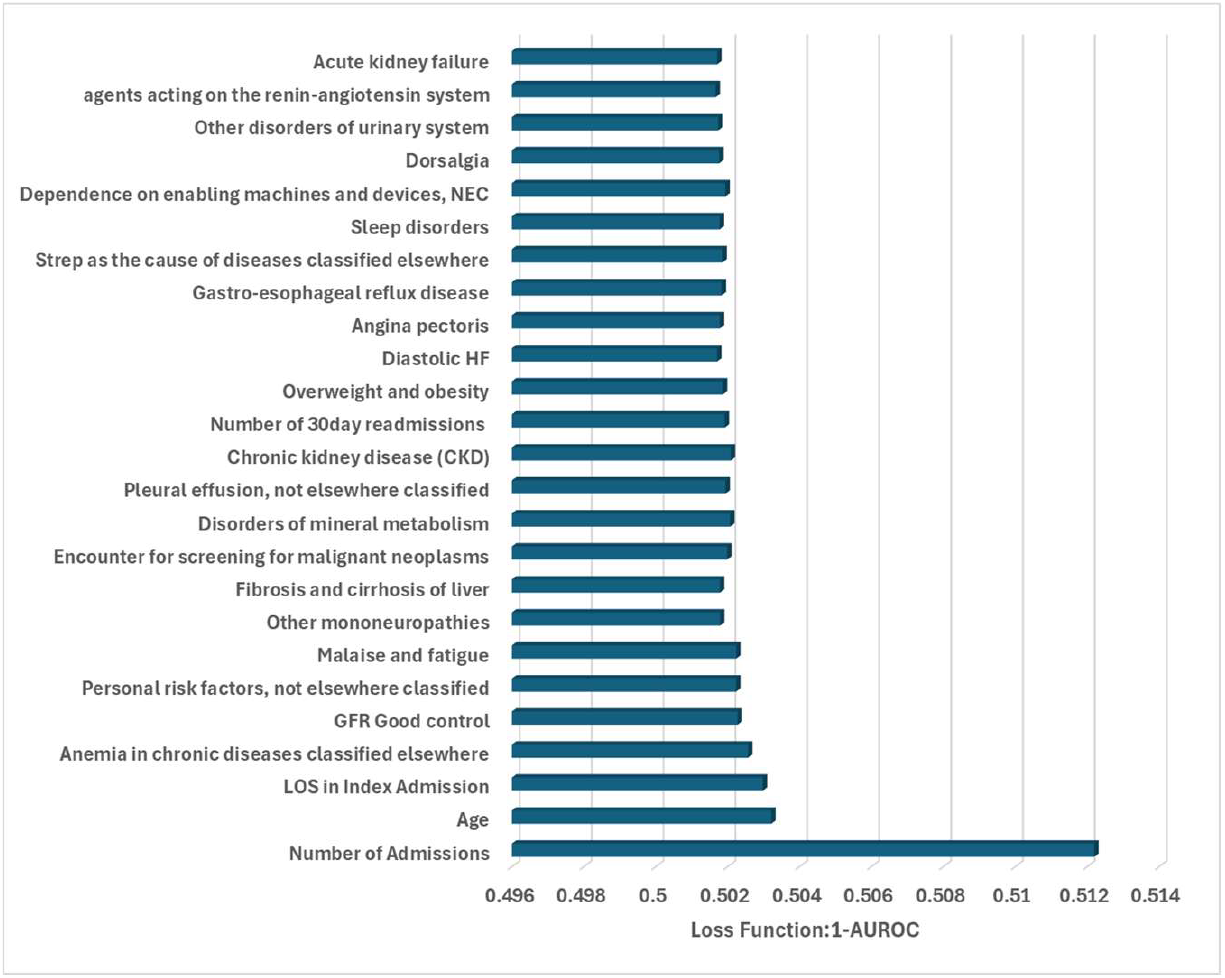
Permutations Feature Importance Plot of the XGBoost Model: Permutation feature importance plot showing the top 25 predictors contributing to the XGBoost model’s performance on the testing dataset. Feature importance was quantified as the mean decrease in model performance (1 − AUROC) following random permutation of each feature, averaged across multiple permutations. Higher values indicate greater contribution to readmission risk prediction.

### Identification of High-Risk Patients and Feature Patterns Among True Positives

Our ML models demonstrated strong predictive performance in identifying HF patients at elevated risk for 30-day readmission. Among the evaluated ML approaches, the XGBoost model showed the strongest performance, identifying nearly one-third of all actual 30-day readmissions in the testing dataset.

To characterize patients most likely to benefit from targeted intervention, we focused on patients in the top 10% predicted probabilities (n = 55) from the testing dataset (n = 548). Within this group, XGBoost identified 42 patients who were readmitted within 30 days, which achieved a true positive rate of 76%. Across all five models, true positive rates in the high-risk decile ranged from 71% to 76%, indicating that approximately three out of four flagged patients were correctly classified.

Descriptive comparisons of feature prevalence among true positive and false positive patients highlighted consistent clinical and utilization patterns associated with correct identification. The most prevalent features among true positives included long-term (current) drug therapy, admission within 1 year before the index admission, type 2 diabetes mellitus, other disorders of fluid, electrolyte, and acid-base balance, and impaired kidney function. These patterns were consistent across models and remained stable when the analysis was expanded to the top 15% risk group (Supplemental Tables 3–8), supporting the robustness of high-risk feature profiles.

## DISCUSSION

HF remains one of the leading causes of 30-day hospital readmissions, often complicated by multiple comorbidities that contribute to both initial hospitalization and re-hospitalization^5,9,29^. Prior research has established that patients admitted with HF as a secondary diagnosis face a similar risk of early readmission as those with a primary HF diagnosis^9,10^. Because HF frequently appears among hospitalization diagnosis codes beyond the primary position, we did not restrict our cohort to primary-diagnosis HF admissions, thereby capturing a broader spectrum of HF-related hospitalizations.

Both RF and XGBoost models demonstrated strong discriminative performance, achieving AUROC values of 0.75 and AUPRC values of approximately 0.58. These results are consistent with established HF readmission models in primary-HF cohorts (AUROC 0.70–0.80) and exceed the performance typically observed in studies that include both primary and secondary HF patients (AUROC 0.62–0.70). Our models achieved true positive rates between 71% and 76% among the top 10% of the risk distribution. Expanding the analysis to the top 15% of high-risk patients further validated the models’ effectiveness, as approximately three out of four patients identified through ML-based risk stratification were correctly classified. Subgroup analyses demonstrated that model performance was stable across sex, age categories, and HF subtypes, with no evidence of systematic performance degradation.

A major strength of our approach is the inclusion of secondary HF encounters, which account for up to 75% of HF-related hospitalizations^9,10^. Because risk factors appear similar across primary and secondary HF, restricting analyses to primary HF omits important clinical profiles and may underestimate true readmission risk. By integrating this broader population, our model identifies risk patterns that are obscured in narrower cohorts, enhancing generalizability and supporting more realistic, real-world clinical application.

Permutation feature importance analysis highlighted prior healthcare utilization, kidney disease and diabetes-related markers, and fluid/electrolyte disorders as key predictors of readmission risk. These findings are clinically intuitive and align with established literature showing that comorbidity burden, acute kidney injury, chronic kidney disease, and prior hospital encounters are strong indicators of future utilization. Patients requiring long-term medication therapy, those with multiple admissions or 30-day readmissions in the year before the index encounter, and those with poor chronic disease management emerged as consistent high-impact predictors across all ML models. The consistency of these predictors across diverse ML approaches reinforces their clinical relevance and suggests they may serve as meaningful targets for intervention.

Mental health conditions were also among the most significant predictors of readmission. These conditions can negatively affect self-care behaviors, medication adherence, and engagement with outpatient follow-ups, thereby increasing the likelihood of hospital readmissions^10,43–48^. This study reinforces the need for proactive management of HF patients with coexisting psychiatric or addiction disorders, including substance abuse and psychosis, to reduce the burden of readmissions.

This study has several limitations that should be considered when interpreting the findings. First, while the KHIN data is extensive, encompassing over 500 healthcare organizations in Kansas and surrounding areas, this cohort was restricted to Kansas residents to ensure robust longitudinal capture of healthcare utilization. As a result, the findings primarily reflect care delivered within the Kansas healthcare system and may not fully generalize to regions with different care delivery patterns or health information exchange participation. In addition, this dataset may overrepresent rural populations and the elderly, and underrepresent urban areas and diverse racial groups, potentially limiting generalizability to other regions. Second, although standardized procedures were applied to address missingness and harmonize diagnostic, medication, and laboratory coding, inherent challenges of EHR-derived HIE data remain, including heterogeneous data formats and variable completeness across contributing sites. Furthermore, KHIN data provide diagnosis codes but do not consistently distinguish between primary and secondary diagnosis positions. As a result, HF hospitalizations could not be stratified by diagnosis hierarchy, which may limit direct comparability with studies relying on primary-diagnosis definitions. Third, the analysis relied exclusively on structured data, which may not capture the depth of clinical nuance present in free-text notes, discharge summaries, or provider narratives; integrating natural language processing in future work may improve predictive accuracy. Collectively, these limitations suggest the need for future external validation using multi-regional or nationally representative datasets to enhance generalizability and evaluate model performance across diverse patient populations.

Despite these limitations, this study demonstrates the strong predictive utility of ML-based models for identifying patients at high risk of 30-day readmission following a HF hospitalization. By leveraging a large, diverse health information exchange dataset, the ML models achieved consistent discrimination, robust calibration, and stable performance across demographic and clinical subgroups. These findings highlight the potential of ML-driven risk stratification to support early identification of vulnerable patients and inform targeted post-discharge interventions aimed at reducing avoidable readmissions. Future work should focus on external validation across multi-regional or nationally representative datasets and on integrating real-time predictive capabilities into clinical workflows to enhance the practical impact of these tools on HF management.

## DECLARATION

### Data Availability Statement

All the clinical data used in this study consist of a limited data set of electronic health record (EHR) data collected by the Kansas Health Information Network (KHIN). The data include a fabricated person identifier and exclude direct personal identifiers in accordance with HIPAA regulations. The dataset was used under a data-use agreement with KHIN and is not publicly available due to privacy and security restrictions.

### Competing Interests

The authors declare that they have no known competing financial interests or personal relationships that could have appeared to influence the work reported in this paper.

### Funding

This research did not receive any specific grant from funding agencies in the public, commercial, or not-for-profit sectors.

### Authors’ Contributions

All the included authors have made significant contributions to this study. All authors reviewed the manuscript, had full access to all data in the study, and took responsibility for the integrity and accuracy of the data analysis and its interpretations. M.K., J.Y., and R.A. wrote the main manuscript. M.K. performed the analysis, and M.K., J.Y., and R.A. led the interpretation of results. All authors contributed to the results and discussion and approved the final version of the manuscript.

### Institutional Review Board Statement

This study was reviewed and approved by the Kansas Health Information Network Data Governance Board and determined to be exempt from institutional review board (IRB) oversight as it involved a secondary analysis of a limited HIPAA dataset lacking identifiable information.

### Informed Consent Statement

Not applicable. This study involved secondary analysis of de-identified and limited data for which informed consent was not required.

## Acknowledgments

The authors thank the Kansas Health Information Network (KHIN) for providing access to the limited dataset used in this research. The authors also acknowledge institutional support for data management and analytic resources that facilitated this study.

## Notes

### Competing Interest Statement

The authors have declared no competing interest.

